# An Analysis of Outbreak Dynamics and Intervention Effects for COVID-19 Transmission in Europe

**DOI:** 10.1101/2020.07.21.20158873

**Authors:** Wei Wang

## Abstract

As of March 13, 2020, Europe became the center of COVID-19 pandemic. In order to prevent further spread and slow down the increase in confirmed cases and deaths, many countries in European Union have taken some interventions since mid-March. In this study, a metapopulation model was used to model the outbreak of COVID-19 in Europe and the effectiveness of these interventions were also estimated. The findings suggested that many countries successfully kept the reproduction number *R*_*t*_ less than 1 (e.g., Belgium, Germany, Spain, and France) while other countries exhibited *R*_*t*_ greater than 1 (e.g., United Kingdom, Cyprus). Based on the assumed reopen strategy, this study also revealed that a 2-week delay in response predicted approximately 2,000 deaths and 200,000 cases (daily peak value), while a 3-week delay predicted approximately 5,000 deaths and 600,000 cases (daily peak value). Therefore, a quick response upon signs of a re-emerging pandemic in the world is highly imperative to mitigate potential loss of life and to keep transmission of Covid-19 under control.

## Introduction

On January 24, 2020, France reported the first case of COVID-19 and by March 17, all European countries had at least one confirmed case when Montenegro confirmed a COVID-19 case [1]. As of March 13, 2020, the World Health Organization (WHO) considered Europe as the epicenter of the COVID-19 pandemic when the number of new reported cases and deaths became greater than those in China [2]. On March 17^th^, the European Union closed all external borders and the next day more than 250 million people were in lockdown across Europe along with already prescribed non-pharmaceutical interventions (NPIs) [3,4].

The NPIs for some European countries were summarized by Flaxman *et al*., (2020) who classified NPIs into five categories: lockdown, public events ban, school closure, self-isolation, and social distancing recommendations [5]. These NPIs were mainly implemented in mid-March 2020 for most European countries and they have been proven successful in controlling the spread of COVID-19 pandemic by reducing *R*_*t*_ < 1 (R_t_ : Effective Reproduction Number, defined as the actual average number of infectious individuals per infection at time t).

Recently, as the number of new reported cases and deaths decreased, many European countries announced relaxation of NPIs and reopening of borders. In this analysis, we aimed to estimate the effectiveness of NPIs in mitigating the spreading of COVID-19 for countries in European Union and to model potential risks associated with lifting NPIs.

## Materials and methods

### Source of Data

The spreading of COVID-19 was modelled through a mobility network of passenger air travel. The first quarter of 2019 data were downloaded from Eurostat on June 12, 2020 [6] and used to describe the inter-country mobility in the transmission model. We drew the COVID-19 outbreak data of all 28 countries of the European Union from the reported confirmed and death cases until June 16, 2020 [7].

### Epidemiology Model

A metapopulation model was used to model the mobility between countries of European Union as described by Ruiyun *et al*., (2020). In this model, besides to the number of people who were S (susceptible), E (exposed), *I*^*r*^ (infected, documented), *I*^*u*^ (infected, undocumented), the parameters were also included in this model as described in Equation 1:

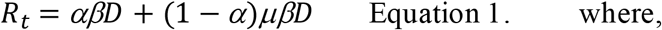

where,

*R*_*t*_ : The effective reproduction number,

β: Transmission rate due to documented infection population,

μ: Reduced factor of transmission rate for undocumented infection population,

α: The fraction of documented infected by the total infected population,

Z: The average latency period, and

D: The average duration of infection.

To calibrate the transmission model against daily incidence and daily death, EAKF (ensemble adjustment Kalman filter) and IF-EAKF (iterated filtering-EAKF) were implemented for different phases of the COVID-19 epidemic [8,9]. In phase one, defined as the time period from February 12, 2020 through March 2, 2020, the IF-EAKF algorithm was used and its parameters were set as *β* ∈ [0.3,1.5], *μ* ∈ [0.2,1.0], *Z* ∈ [2,5], *D* ∈ [2,5], *α* ∈ [0.02,1.0], *θ* ∈ [0.01,10.0]. In phase two, defined as the time period from March 3, 2020 through June 16, 2020, the EAKF algorithm was implemented and its parameters were set as *β* ∈ [0.05,1.5], *μ* ∈ [0.1,1.0], *Z* ∈ [2,5], *D* ∈ [2,5], *α* ∈ [0.05,1.0], *θ* ∈ [0.01,1.0]. As there were many outbreaks at the same time for European countries, the model was initiated by using the same strategies described by Pei Sen *et al*., 2020. *E* and *I*^*u*^ were randomly drawn from uniform distributions [0,12*C*] and [0,10*C*] eight days prior to the reporting date (*T*_0_) of the first case. Here, *C* was the total number of reported cases between *T*_0_ and *T*_0_ + 4.

### Reopening Strategy

The control measures were further relaxed on June 17, 2020 for all 28 countries.

*R*_*t*_ for all countries were maintained with values higher than 1.5. For countries with *R*_*t*_ values lower than 1.5, the corresponding values were set as 1.5. The daily incidence and daily death for six weeks were projected using the calibrated models and after a response time of 2 or 3 weeks, a 25% weekly reduction of transmission rates were assumed.

## Results and Discussion

As seen in Table 1, the inferred parameters (±95% CI) were reported for the best-fit model ranging from February 12, 2020 through March 2, 2020. The results showed that only 6% of total infections were reported during this period. A high proportion of infection population (94%) was not reported because of mild or asymptomatic infections or due to low detection rate. Our result showed that those undocumented infections were contributed to the COVID-19 pandemic with approximately half the level of contagiousness (α=0.52; 95%CI: 0.46, 0.58) which is similar to the result reported by Ruiyun *et al*., (2020). Owing to no symptoms and poor prevention awareness, most undocumented infections do not seek medical care. Based on the reasons mentioned above, those undocumented infections facilitate the rapid spread of COVID-19.

**Table 1.**
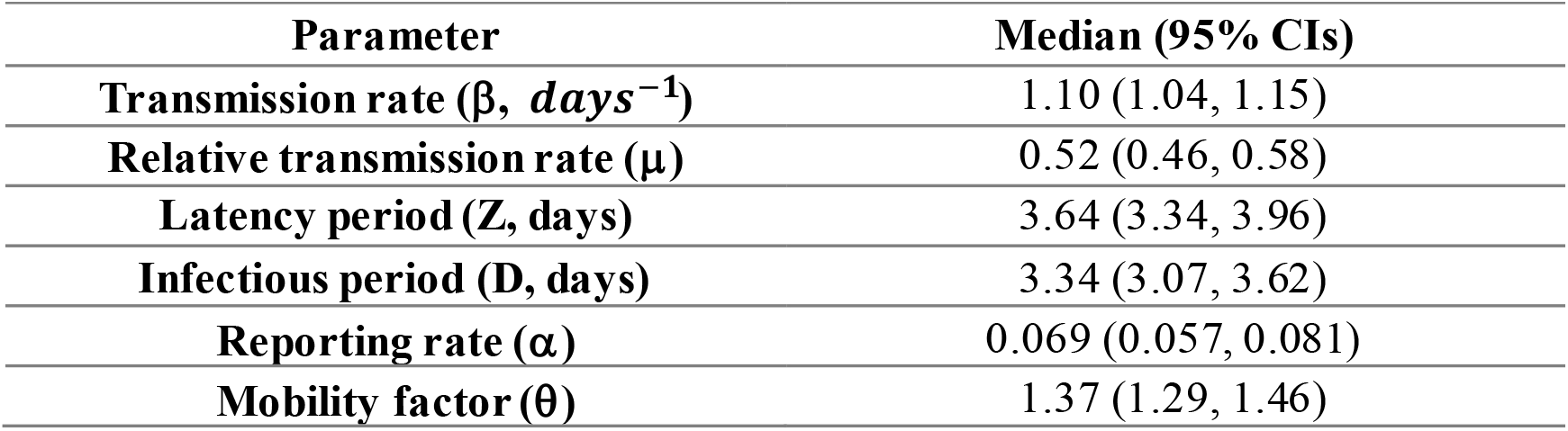
Best-fit model posterior estimates of key epidemiological parameters

After March 2, 2020, many NPIs were implemented in European countries at different dates. Therefore, the same β was not used for all 28 countries. The EAKF algorithm was used to fit the daily incidence and death to estimate the R_t_ values for different countries (Figure 1). As seen in Table 2, many countries successfully kept the *R*_*t*_ value less than 1, (e.g., Belgium, Germany, Spain, France) while other countries exhibited *R*_*t*_ greater than 1 (*R*_*t*_ > 1), (e.g., United Kingdom, Cyprus), indicating insufficient control of the pandemic.

**Table 2.**
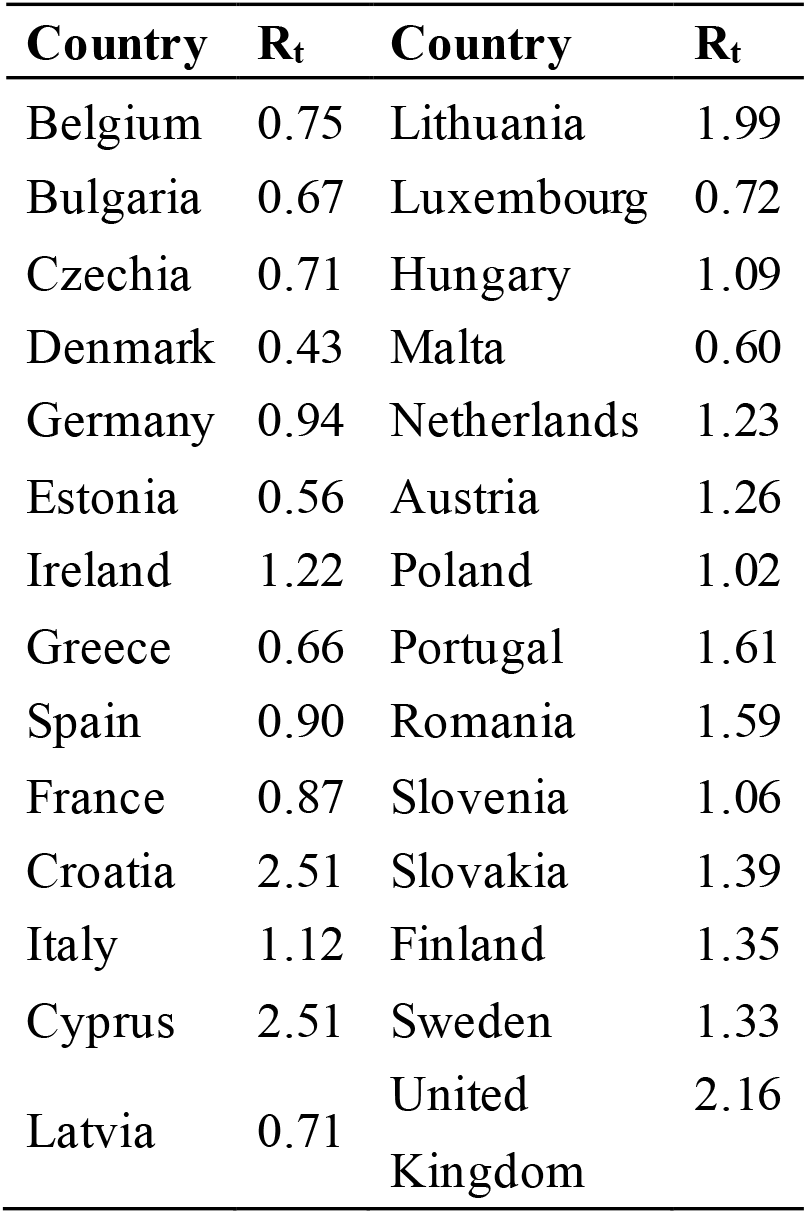
*R*_*t*_ values for 28 countries on June 8, 2020

**Figure 1.**
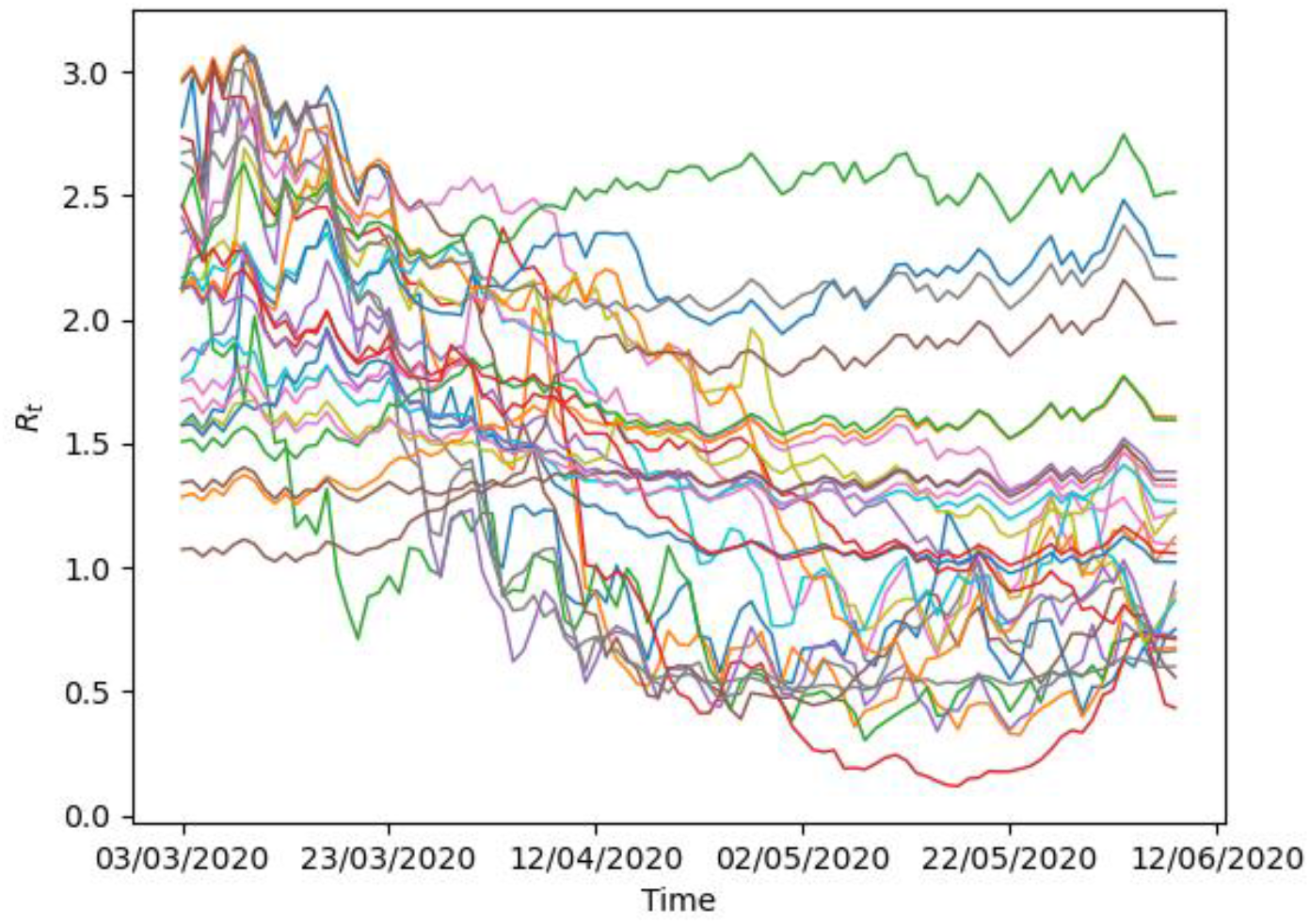
R_t_ value change from March 3, 2020 through June 8, 2020 for different countries (Country Names are not shown). The EAKF algorithm was used to fit the daily incidence and death to estimate the R_t_ values for different countries.

The estimated European ascertainment rate was declined from 0.2 to 0.06 in mid-April with a rapid COVID-19 spread indicating that the increase of testing capacity was insufficient to keep up with the increase in infections (Figure 2). Once there was an observable decrease in cases in mid-April with increased testing capacity, the ascertainment rate was increased back to its earlier levels (∼0.2).

**Figure 2.**
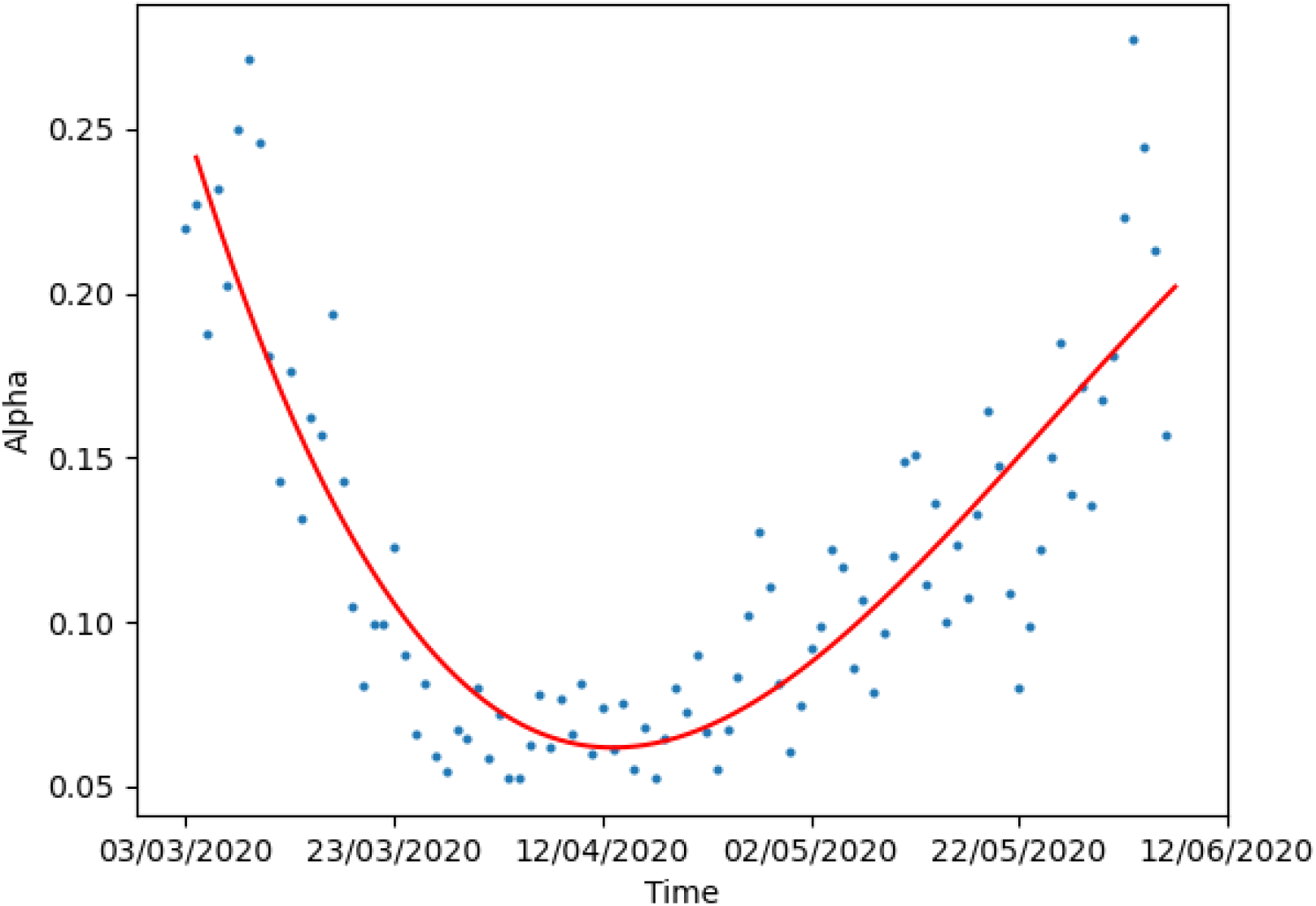
The estimated ascertainment rate α over time. Dots show the estimated α values and the red line is the fitted polynomial line.

Furthermore, reopening and potential risks were also investigated. In this study, an assumed NPI relaxation was modelled on June 17, 2020 for potential effects on infections and deaths in these European countries. In these instances, the R_t_ was anticipated to be ≥1.5 for all countries, depending on the R_t_ prior to June 17^th^. The difference in COVID-19 impacts was specifically investigated if NPIs with 25% efficiency were implemented each week following either a 2 - or 3-week initial response time delay. A 3-week response time to re-implement NPIs predicted large numbers of confirmed infections and deaths (Figure 3). In the meantime, a 2 -week delay predicted approximately 2,000 deaths and 200,000 cases (daily peak death), while a 3-week delay predicted approximately 5,000 deaths and 600,000 cases (daily peak death). Therefore, quick action upon signs of a re-emerging pandemic in these countries was imperative to mitigate potential loss of life. Finally, it should be noted that this model would support implementation of the most effective NPIs very early in an effort to reduce R_t_ < 1 in these countries.

**Figure 3.**
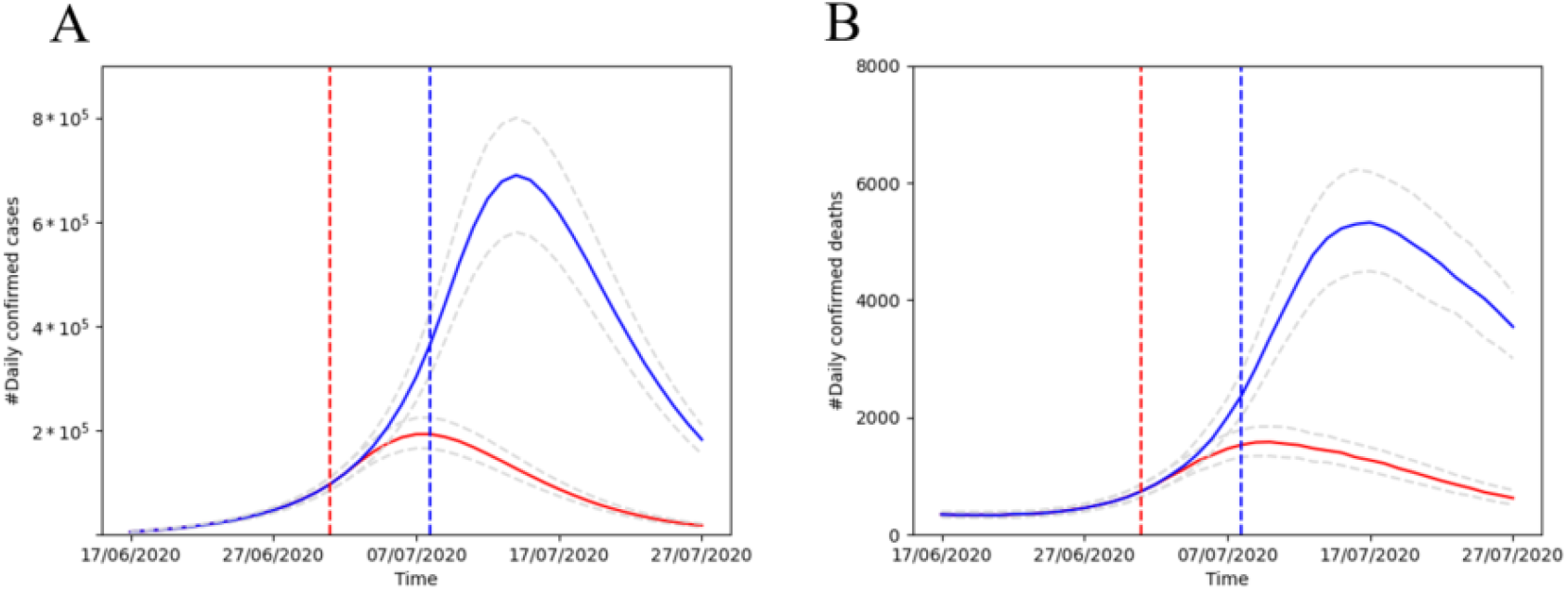
NPIs are further relaxed after June 17, 2020 in all European countries. (A) daily confirmed cases after re-open for 2 weeks (red line) and 3 weeks (blue line). The dash lines show the 25%-75% CIs. (B) daily confirmed deaths after re-open for 2 weeks (red line) and 3 weeks (blue line). The dash lines show the 25%-75% CIs.

## Data Availability

No

